# Radiologist Performance and Reliability of MR Scoring Systems in Perianal Fistulizing Crohn’s Disease: Retrospective Analysis in a Real-World Clinical Practice

**DOI:** 10.1101/2023.10.07.23296649

**Authors:** Suha Abushamma, Ravella Balakrishna, Alvin George, John Hickman, Daniel R. Ludwig, Anup S. Shetty, Maria Zulfiqar, Aravinda Ganapathy, Grace Bishop, Anusha Elumalai, Jyoti Arora, Philip Miller, Parakkal Deepak, David H. Ballard

**Affiliations:** Division of Gastroenterology, Washington University School of Medicine in St. Louis, St. Louis, MO, USA; Department of Gastroenterology & Hepatology, Cleveland Clinic, Cleveland, OH, USA; Division of Gastroenterology, University of Virginia, Charlottesville, VA, USA; Mallinckrodt Institute of Radiology, Washington University School of Medicine in St. Louis, St. Louis, MO, USA; Department of Radiology, Mayo Clinic, Scottsdale, AZ, USA; Institute for Informatics, Data Science & Biostatistics, Washington University School of Medicine in St. Louis, St. Louis, MO, USA

**Keywords:** perianal fistula, anal fistula, perianal Crohn’s disease, Crohn’s disease, inflammatory bowel disease, magnetic resonance imaging

## Abstract

**Background:** Perianal fistulizing Crohn’s disease (CD-PAF) is difficult to treat, and several MR scoring systems have been developed to assess treatment response.

**Objective:** To assess the relationship between CD-PAF MR scoring systems and radiologists’ subjective assessment of CD-PAF severity and treatment response on baseline and follow-up pelvic MR.

**Methods:** Retrospective single institution study of consecutive symptomatic patients with CD-PAF patients who underwent pelvic MR before and ≥3 months after initiating biologic therapy during a 10-year period (December 2011 to December 2021). One of four radiologists interpreted baseline and follow-up MRs. Scoring systems included variables in the modified Van Assche index (mVAI), magnetic resonance novel index for fistula imaging in CD (MAGNIFI-CD), and pediatric MR-based perianal Crohn’s disease (PEMPAC) index. For initial and follow-up MR, a 5-point Likert scale assessed severity (1=mild, 3=moderate, 5=marked). On follow-up MR, radiologists evaluated fistula response as 1-improved, 3-stable, or 5-worsened CD-PAF severity. All four study radiologists scored the baseline MR in 20 patients to calculate inter-reader agreement statistics. Interrater reliability was assessed with the Krippendorff α coefficient for categorical variables and intraclass correlation coefficient (ICC) for continuous variables.

**Results:** The final cohort included 96 CD-PAF patients (50 men; mean age=33.0 years) with 192 baseline and follow-up MRs. Moderate to substantial agreement was observed among radiologists for MAGNIFI-CD, mVAI, PEMPAC, and Likert scores (ICC: 0.716, 0.756, 0.535, and 0.679 respectively). Individual components of MR scoring systems had Fair to Substantial agreement (Alpha: 0.195 to 0.730). Significant univariate associations were found between MR scoring systems and radiologists’ Likert severity assessments (p<0.001, Pearson correlation coefficients ≥0.820). In patients meeting criteria for change in disease severity (n=17), all scoring systems demonstrated AUC values ≥0.93.

**Conclusion:** The MR scoring systems for CD-PAF (MAGNIFI-CD, mVAI, and PEMPAC) demonstrated strong associations with radiologists’ subjective assessments of severity and treatment response on baseline and follow-up pelvic MR. Inter-reader agreement of these scoring systems outperformed individual MR factors.

## Introduction

Crohn’s disease (CD) is a chronic, progressive inflammatory disorder of the intestinal tract that affects up to 1% of the population in Western countries [1,2]. Population-based studies estimate that perianal fistulas develop in approximately 13% – 27% of Crohn’s disease (CD) patients, with the cumulative risk for developing a perianal fistula being 21% after 10 years and 26% – 28% after 20 years [3].

Magnetic resonance imaging (MRI) of the pelvis is the standard of care in assessment of perianal fistulizing Crohn’s disease (CD-PAF) [4]. MR interpretations of perianal fistulas are pivotal in guiding and evaluating the response to medical management and surgical planning [5]. Furthermore, radiological healing or remission beyond clinical cessation of drainage is a recent concept [6,7] in managing CD-PAF where fistula tracts that are persistent on MR despite apparent clinical closure [8] have higher relapse rates [9] and need for therapeutic interventions [10]. In contrast, radiological healing with fibrosis of the fistula tract leads to sustained fistula resolution, regardless of whether anti-TNF therapy is continued or stopped [6, 11, 12].

Several MR pelvis-based imaging scoring systems have been developed and validated using several imaging variables [4]. However, the utility of these scores in real-world cohorts is unclear. The purpose of our study was to assess the relationship of CD-PAF MR scoring systems to radiologists’ subjective assessment of CD-PAF severity and treatment response on baseline and follow-up pelvic MR.

## Materials and Methods

In a retrospective design, we screened consecutive symptomatic patients with CD-PAF seen at our center from December 1, 2011, to December 31, 2021, who underwent fistula protocol MR of the pelvis before initiating biologic therapy for CD-PAF. The protocol and conduct of this study conformed to the 1975 Declaration of Helsinki and the Health Insurance Portability and Accountability Act. Institutional review board approval was obtained (local IRB #202004277), including a waiver of informed consent. Inclusion criteria were symptomatic CD-PAF (perianal symptoms included drainage, pain, or pruritus at or surrounding the anal canal), inpatient or outpatient consultation by one of our board-certified gastroenterologists, a diagnostic MR of the pelvis before the initiation of a new medical treatment for CD-PAF, and a follow-up MR of the pelvis at least 3 months after the initiation of medical treatment of CD-PAF. A CD-PAF diagnosis was qualified if they were diagnosed by a board-certified gastroenterologist practicing at one of our center’s Inflammatory Bowel Disease clinics. A qualifying diagnostic MR was a fistula protocol MR of the pelvis with adequate small field-of-view high-resolution T2-weighted sequences, as determined by one of the study radiologists to accurately characterize perianal fistula.

In our practice, MR pelvis fistula protocol without and with contrast is performed in patients with suspected or known CD-PAF, often performed in conjunction with an MR enterography examination of the abdomen. Follow-up examinations are not standardized for symptomatic CD-PAF and are determined by the managing gastroenterologist. In asymptomatic CD-PAF patients or patients with CD-PAF who achieve improved symptoms, follow-up imaging is usually obtained at 1-year intervals. Exclusion criteria included lack of a clear CD diagnosis, biologic therapy at time of initial CD-PAF evaluation, lack of an MR with adequate diagnostic sequences (determined during study imaging review by study radiologists), and history of proctectomy at time of initial MR (fecal diversion without proctectomy was not an exclusion criterion).

Potentially eligible patients who met all inclusion criteria except for radiologist evaluation of the adequacy of the MR pelvis fistula protocol underwent a structured chart review for demographic information, medical and surgical treatment of CD-PAF, and follow-up data. Data was entered into our institutional Research Electronic Data Capture (REDCap) system [13].

### Imaging analysis

#### MR protocol

MR pelvis fistula protocol was performed without and with contrast on 1.5-T or 3.0-T Siemens (Erlangen, Germany) MR scanners. MR pelvis fistula protocol included a small field-of-view high-resolution T2-weighted turbo spin echo through the anal canal along with T2, pre-contrast T1, and post-contrast T1 through the pelvis. Contrast sequences through the pelvis were different when examinations were MR pelvis fistula protocol and MR enterography paired with MR pelvis fistula protocol. When a dedicated MR pelvis fistula protocol was performed, dynamic contrast sequences (arterial, venous, and delayed phases) were obtained through the pelvis. When fistula protocol pelvis sequences were performed concurrently with MR enterography, dynamic contrast sequences (arterial, venous, and delayed phases) were obtained through the abdomen and small bowel, and a single pelvis post-contrast acquisition was obtained in a delayed phase.

#### Training

Image evaluation was performed by one of four board-certified abdominal radiologists with fellowship training with 1-, 3-, 4-, and 6-years post-fellowship who regularly interpret MR pelvis fistula protocol in CD-PAF as part of their clinical practice. The four study radiologists met for a 1.5-hour training session where the definitions of the MR scoring systems were reviewed in a training set of sample cases not in the imaging cohort of potentially eligible patients. After discussing variables not accounted for in the MR scoring systems, fistula extension to the genitals (scrotum, penis, labia, or vagina) was added as a study variable. The study radiologists met before the imaging review for the initial training session and met a second time after each radiologist reviewed the same 5 patients. The second meeting was performed to resolve any unforeseen issues in imaging interpretation, which included how to record T1-hyperintensity in patients without dynamic contrast acquisitions through the pelvis and fistula length measurements. As noted, our institutional protocol differs for contrast acquisitions when MR pelvis fistula protocol is performed concurrently with MR enterography, and MAGNIFI-CD requires assessment of T1 hyperintensity. The option for the readers to mark “not assessable” for T1 hyperintensity was added to account for this. The other issue addressed at the second radiologist meeting was agreeing on a standard method to measure fistula length. A standard fistula measurement was determined, adding linear measurements on a single sequence that the radiologist thought best showed the fistula extent.

#### Study imaging interpretation

MR scoring systems included variables in the modified Van Assche index (mVAI) [14], magnetic resonance novel index for fistula imaging in CD (MAGNIFI-CD) [15], and pediatric MR-based perianal Crohn disease (PEMPAC) index [16]. A formal calculation of the original Van Assche index (VAI) was not performed [17]. The study radiologists recorded the components of the three CD-PAF MR scoring systems along with fistula extension to the genitals (scrotum, penis, labia, or vagina). For initial and follow-up MR evaluation, a 5-point Likert scale assessment of subjective severity was established where 1 indicated mild severity, 2-mild to moderate severity, 3-moderate severity, 4-moderate to marked severity, and 5-marked severity. When assessing fistula response on follow-up MR, we asked the study radiologists to rank the MR imaging findings that led to their decision to mark it on the Likert scale as 1-substantially improved, 2-mildly improved, 3-stable, 4-mildly worsened, or 5-substantially worsened CD-PAF severity. We intently chose not to have formal definitions for the Likert scale severity or Likert scale radiological response at follow-up. Study radiologists were instructed to use the Likert scores as they would if they chose to use these severity descriptors in a clinical interpretation. For example, a clinical interpretation of: i) moderate to marked perianal CD with two complex transsphincteric perianal fistulas feeding a small abscess in the left ischioanal fossa may translate to a 4- or 5-point severity; ii) this appears unchanged to slightly improved from the prior MR 8 months earlier would translate to a 2- or 3-point follow-up Likert scale.

The study radiologists were blinded to all clinical information aside from patient age and sex. They had access to both initial and follow-up MR examinations and were instructed to finalize their score for the initial MR before proceeding to the follow-up MR. When inputting the components of each MR scoring system, the numeric score was not calculated. Another study investigator not involved in imaging interpretation later calculated the numeric scores of each MR scoring system. Image interpretation mirrored radiologists’ clinical workflow and practice, performed on a clinical workstation using the Sectra picture and archiving communication system (Sectra PACS Workstation IDS7, Sectra AB, Linköping, Sweden). Each study radiologist read 18 unique patient initial and follow-up MR (36 MR examinations). For inter-reader assessment, all four study radiologists read 20 patients’ initial MR examinations along with 5 patients’ initial and follow-up MR examinations (30 MR examinations in 25 patients for inter-reader assessment).

### Statistical analysis

The primary statistical analyses focused on the responsiveness of each MR scoring system against radiologist assessment of initial and follow-up MR pelvis disease severity and response using the Likert scale to detect a 2-point or greater change in Likert score. Pearson correlation coefficients were calculated to measure the association between the baseline and follow-up MR scores and the Likert severity scores assigned by the study radiologists. The non-parametric Kruskal-Wallis test was employed to determine the p-values for the univariate association analysis between the MR scoring systems and the study radiologists’ subjective overall impression. Quantitative variables with a normal distribution are presented as means ± SDs. Medians and interquartile ranges (IQRs) were adopted for those variables not following a normal distribution. Categorical variables are detailed in terms of counts and percentages.

Interreader agreement was assessed using the Krippendorff α coefficient, a weighted reliability measurement tool with values ranging from 0 (indicating no agreement) to 1 (signifying complete agreement). A Krippendorff α value below zero suggests systematic disagreements among readers instead of random chance. The Krippendorff α values for each imaging feature and bootstrap 95% confidence intervals were computed using the **icr** package [18] in R software to gauge the consistency and reliability of the radiologists’ evaluations. Krippendorff’s alpha is versatile and can handle data from multiple raters, while Cohen’s kappa is primarily designed for two raters. Another feature of Krippendorff’s alpha is its adaptability to different levels of measurement, such as nominal, ordinal, interval, and ratio, giving it an edge in flexibility over kappa statistics. This analysis used ordinal weighting to capture the severity or magnitude of disagreement amongst radiologists. Krippendorff’s alpha offers an advantage in handling instances of missing data and operates under fewer restrictive assumptions compared to kappa statistics [19].

Intraclass correlation coefficient (ICC) was used for the continuous variables of fistula size measurements and MR scoring systems. Agreement was scored as poor (α< 0.00), slight (α = 0.00–0.20), fair (α = 0.21–0.40), moderate (α = 0.41–0.60), substantial (α = 0.61–0.80), or almost perfect (α = 0.81–1.00) [20].

## Results

### Study Cohort

Of 98 patients evaluated by study radiologists, 2 patients were excluded, 1 with inadequate initial imaging for fistula characterization and 1 with proctectomy at initial MR that was not appreciated at the initial chart review. 96 CD-PAF patients with 192 baseline and follow-up MRs constituted the final study cohort. The 96 patients included 50 men and 46 women with a mean age of 33.0 years (SD +/-16.1 years) at baseline MR (**Table 1**).

**Table 1.** Baseline characteristics of 96 perianal Crohn’s disease patients included in this study.

| **Variable** |  | **N = 96** | **%** |
| --- | --- | --- | --- |
| **Sex** | Male | 51 | 53.13 |
|  | Female | 45 | 46.88 |
| **Age at Diagnosis (Mean)** |  | 24.27 (+/-14.79) | - |
| **Age at Baseline MR (Mean)** |  | 32.95(+/-16.13) | - |
| **Disease Duration in Years (Mean)** |  | 8.68(+/-9.90) | - |
| **Baseline Prior Medication** | No | 27 | 28.13 |
|  | Yes | 69 | 71.88 |
| **Montreal Age Classification at Time of Diagnosis** | 16 years or younger (A1) | 39 | 40.63 |
|  | 17-40 years of age (A2) | 44 | 45.83 |
|  | >40 years of age (A3) | 13 | 13.54 |
| **Montreal behavior classification** | Nonstricturing, nonpenetrating (B1) | 18 | 18.75 |
|  | Stricturing without penetrating disease (B2) | 3 | 3.13 |
|  | Penetrating disease (B3) | 75 | 78.13 |

### MR Evaluation and Scoring Systems

Three MR scoring systems, MAGNIFI-CD, mVAI, and PEMPAC, were scored by at least one study radiologist for all 96 patients, along with a 1 to 5 Likert severity score (with 5 being the most severe). 85 patients had complete data for both initial and follow-up MAGNIFI-CD scores, 95 had complete data for both initial and follow-up mVAI scores, 96 had complete data for both initial and follow-up PEMPAC scores and all 96 patients had complete data for both initial and follow-up subjective Likert severity scores. Eleven patients did not have adequate T1 postcontrast sequences on their initial or follow-up MR to calculate the MAGNIFI-CD score. Mean baseline and follow-up MR scores and Likert severity are presented in **Table 2**. Case examples demonstrating baseline and follow-up MR with applying the study-evaluated MR scoring systems are demonstrated in **Figure 1** and **Figure 2**.

**Figure 1.**
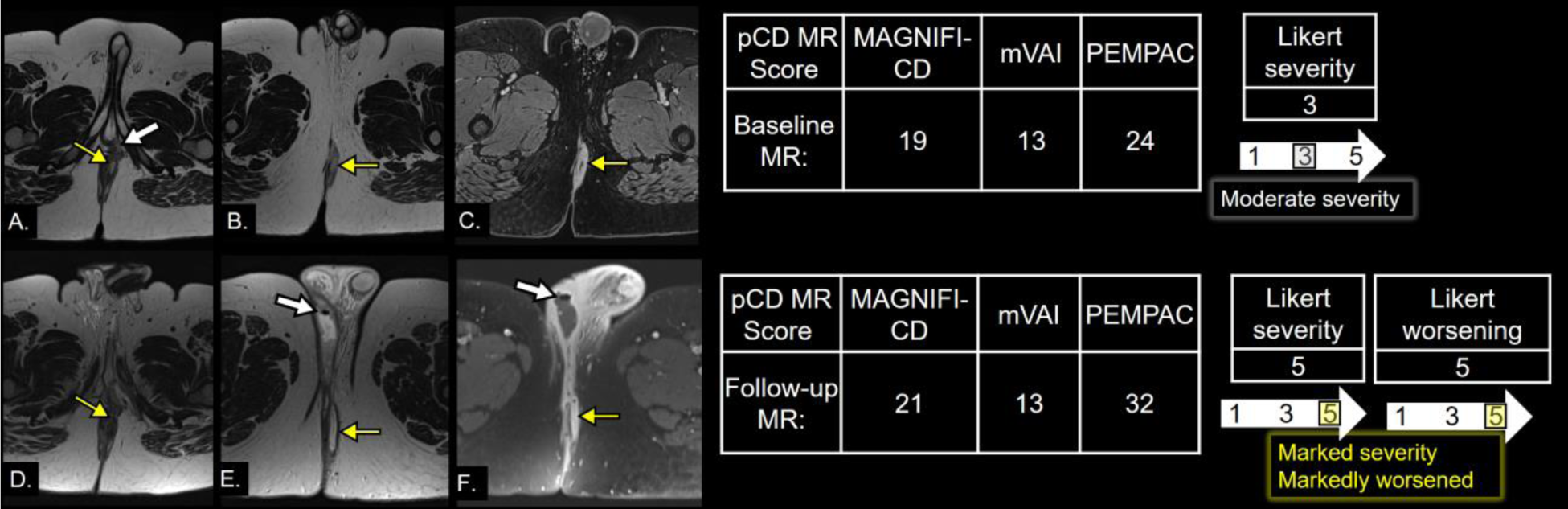
Worsening disease at follow-up MR. Young adult man with perianal Crohn’s disease. **Top row, A-C**. Initial baseline MR pelvis fistula protocol without and with contrast with axial small field-of-view high-resolution T2 turbo spin echo axial images at the level of the anal verge (A) and left perineum (B) and an axial T1 postcontrast fat-saturated acquisition at the level of the left perineum (C). A left perineal abscess (yellow arrow in B and C) is fed by an intersphincteric fistula near the anal verge originating from a defect in the internal sphincter at the 1 o’clock position (yellow arrow in A). Inflammation from the fistula tract abuts the base of the penis on the left (thick white arrow in A). Perianal Crohn’s disease MR scores are presented for the baseline MR, and the study radiologist graded this as a 3 of 5 Likert scale severity, corresponding to moderate severity. **Bottom row, D-F**. 6-month follow-up MR acquisition with axial small field-of-view high-resolution T2 turbo spin echo axial images at the level of the anal verge (D) and perineum/scrotum (E) and axial T1 postcontrast fat-saturated acquisition at the level of the low perineum/scrotum (F). The intersphincteric fistula and defect in the internal sphincter near the anal verge are smaller with a less discrete tract (yellow arrow in D); however, the left perineal abscess is larger and tracks more posterior and anterior with a component forming a large abscess in the right hemiscrotum containing locules of gas (thick white arrows pointing to gas in the right scrotal abscess in E and F). MR scores worsened as presented except for the mVAI, the MAGNIFI-CD, and PEMPAC scoring systems had longer fistula length to account for higher scores. The study radiologist noted this is the highest subjective Likert severity, 5 out of 5, markedly severe, and the worst possible change from comparison MR, 5 out of 5, corresponding to markedly worsened perianal Crohn’s disease.

**Figure 2.**
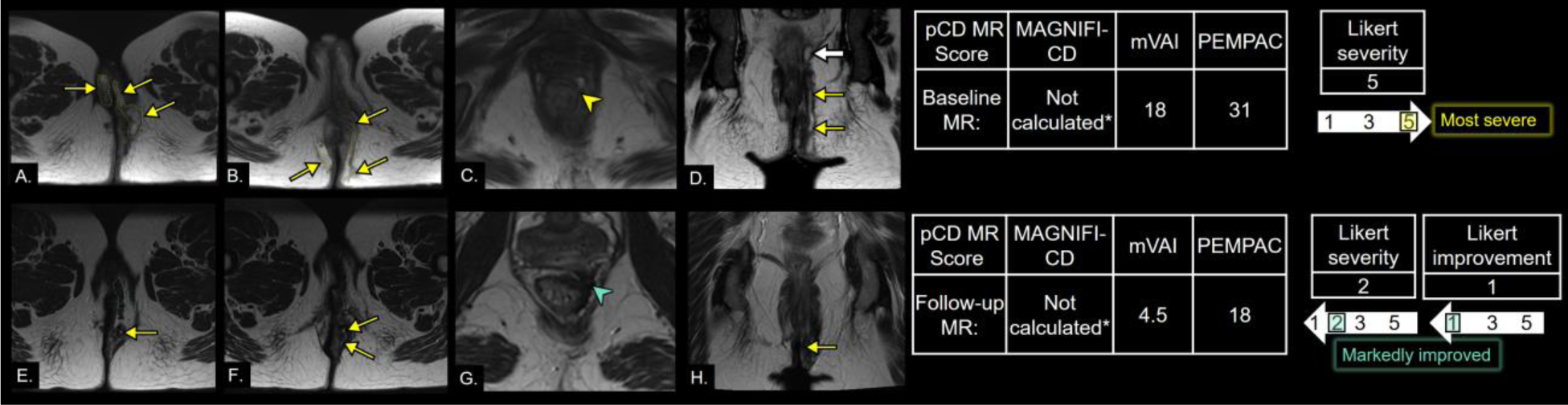
Improving disease at follow-up MR. Young adult woman with perianal Crohn’s disease. **Top row, A-D**. Initial baseline MR enterography with additional fistula pelvis sequences without and with contrast with axial small field-of-view high-resolution T2 turbo spin echo axial images at the level of the perineum/vulva (A), anal verge (B), a cropped in view of the internal sphincter near the anorectal junction (C), and a coronal acquisition through the internal and external sphincter (D). Abscesses fed from a transsphincteric fistula involve the bilateral vulva, left perineum, and bilateral gluteal cleft (yellow arrows in A, B, and D) and the faint dashed yellow outline in A and B). This transsphincteric fistula feeding abscesses originates from a 1:00 defect in the internal sphincter near the anorectal junction (yellow arrowhead in C). The supralevator extension of this complex fistula forms a small abscess adjacent to the distal rectum (thick white arrow in D.). MR scores are presented. The MAGNIFI-CD could not be calculated because the MR enterography contrast acquisition had its early contrast phases focused on the abdomen/small bowel, and T1 hyperintensity is a required component of MAGNIFI-CD. The study radiologist graded this 5 out of 5 on the subjective Likert scale, the most severe category. **Bottom row, D-F**. 8-month follow-up MR acquisition with axial small field-of-view high-resolution T2 turbo spin echo axial images at the level of the perineum/vulva (E), anal verge (F), cropped in view of the internal sphincter near the anorectal junction (G), and a coronal acquisition through the internal and external sphincter (H). There has been interval resolution of abscesses with only a small patent appearing fistulous tract in the left perineum and left gluteal cleft fat (yellow arrows in E, F, and H). There is scar tissue delineated by T2 hypointense signal at the left perineum and vulva (faint dashed blue outline in E), and there is similar scarring without patent fistula at the prior site of internal sphincter opening 1 o’clock position near the anorectal junction (blue arrowhead in G). MR scores improved as presented. The study radiologist noted this as a 2 out of 5 on a Likert severity score and scored it as markedly improved on the Likert follow-up score, corresponding to markedly improved perianal Crohn’s disease.

**Table 2.** Median baseline and follow-up MR scores for 96 perianal Crohn’s disease patients as rated by the study radiologists.

|  | **MAGNIFI-CD (n=85\*)** | **mVAI (n=95)** | **PEMPAC (n=96)** | **Likert severity (n=96)** |
| --- | --- | --- | --- | --- |
|  | **Range=0-25** | **Range=0-19.5** | **Range=0-41** | **Range=0-5** |
| Baseline MR: Median (Q1, Q3) | **14 (10,18)** | **8.2 (4.7,11.4)** | **26 (15,31)** | **3 (2,4)** |
| Follow-up MR: Median (Q1, Q3) | **14 (8,18)** | **6.2 (3.5,11.2)** | **24 (14,29)** | **2 (1,4)** |

\* - 11 patients could not have their initial MAGNIFI-CD calculated due to inadequate T1 postcontrast pelvis coverage with paired MR enterography and fistula pelvis MR examinations.

### Inter-reader Reliability and Agreement

Inter-rater reliability analyses are summarized in **Table 3** and **Table 4**. The baseline MR score showed moderate to substantial agreement among the study radiologists for MAGNIFI-CD, mVAI, and PEMPAC (**Table 3**).

**Table 3.** Inter-reader reliability perianal Crohn’s disease MR scoring systems and length of fistula tract. Intraclass correlation coefficients (ICC) of 4 fellowship-trained abdominal radiologists assigning perianal Crohn’s disease MR scores at baseline and follow-up MR.

| **CD-PAF MR Scoring System** | **ICC Baseline MR (n=24)** | **ICC Follow-up MR (n=5)** | **Agreement Interpretation** |
| --- | --- | --- | --- |
| **MAGNIFI-CD** | 0.716 (0.535-0.846) | 0.497 (0.123-0.875) | Substantial/Moderate |
| **mVAI** | 0.756 (0.602-0.864) | 0.752 (0.359-0.943) | Substantial/Substantial |
| **PEMPAC** | 0.535 (0.339-0.721) | 0.582 (0.181-0.898) | Moderate/Moderate |
| **Length of Fistula Tract** | 0.362 (0.162-0.594) | 0.173 (-0.008 -0.7) | Moderate/Poor |

**Table 4.** Inter-reader reliability individual components of perianal Crohn’s disease MR scoring systems.

| **MR Imaging Component** | **Alpha Baseline MR (n=24)** | **Alpha Follow-up MR (n=5)** | **Agreement Interpretation** | **mVAI weight** | **MAGNIFI-CD weight** | **PEMPAC weight** |
| --- | --- | --- | --- | --- | --- | --- |
| **Number of fistula tracts** | 0.345 (0.163-0.510) | 0.827 (0.633-0.957) | Fair/Almost Perfect | 0% | 24.0% | 19.5% |
| **Location** | 0.240 (0.058-0.412) | -0.114 (-0.739-0.503) | Slight/Poor | 0% | 0% | 29.3% |
| **Extension** | 0.547 (0.443-0.643) | 0.302 (-0.647-0.516) | Moderate/Fair | 23.1% | 16.0% | 0% |
| **T2 hyperintensity** | 0.414 (0.062-0.707) | 0.294 (-0.118 - 1) | Moderate/Fair | 23.6% | 0% | 9.8% |
| **T1 hyperintensity** | 0.195 (-0.058-0.432) | -0.268 (-0.689-0.156) | Fair/Poor | 0% | 8% | 0% |
| **Presence of proctitis** | 0.344 (0.186-0.495) | 1.0 (1 -1) | Fair/Perfect | 10.3% | 0% | 0% |
| **Abscess** | 0.466 (0.326-0.594) | 0.249 (-0.09 – 0.561) | Moderate/Fair | 30.8% | 20.0% | 26.8% |
| **Dominant feature** | 0.730 (0.505-0.91) | 0.220 (-0.341-0.665) | Substantial/Fair | 12.3% | 16.0% | 0% |
| **Likert severity** | 0.679 (0.577 – 0.763) | 0.004 (-0.596–0.462) | Moderate/Poor |  |  |  |
|  | **ICC Baseline MR (n=24)** | **ICC Follow-up MR (n=5)** |  |  |  |  |
| **Length of Fistula Tract** | 0.362 (0.162-0.594) | 0.173 (-0.008 -0.7) | Moderate/Poor | 0% | 16.0% | 15.0% |

For the baseline MR comprising 24 samples (**Table 4**), the agreement ranged from “Slight” for the location component (α=0.240) to “Substantial” for the dominant feature (α=0.730). Notably, evaluating the number of fistula tracts and the presence of proctitis produced “Fair” agreement with α values of 0.345 and 0.344, respectively. Evaluating the number of fistula tracts achieved an “Almost Perfect” agreement with an α value of 0.827. In stark contrast, the location and T1 hyperintensity components recorded negative alpha values, suggesting a “Poor” agreement, with α values of −0.114 and - 0.268, respectively. Most notably, the assessment for the presence of proctitis showed “Perfect” agreement with a Krippendorff’s alpha of 1.0. These results reflect diverse reliability across radiological findings in perianal Crohn’s disease across two different time points. The agreement demonstrated a broader range in the follow-up MR cases, which consisted of 5 patients. Data from follow-up MR agreement should be interpreted with discernment that it was only in a subset of 5 of the 24 patients. The follow-up examinations were part of the evaluated MR data, but we did not design the study to determine inter-reader agreement at follow-up examinations.

### Responsiveness of MR Scoring Systems

The responsiveness of the MAGNIFI-CD, mVAI, and PEMPAC MR scoring systems was assessed by comparing baseline and follow-up MR scores to study radiologists’ Likert severity scores and assessment of changes at follow-up. Univariate association of MR scoring systems with study radiologists’ subjective Likert disease severity showed significant (p<0.001 in all analyses) Pearson correlation coefficients of 0.820 or higher (**Table 5**).

**Table 5.** Univariate association of MR scoring systems with subjective severity by Likert score (1-5) assigned by the study radiologists.

| **Variable vs. Likert Disease Severity** | **Pearson CC** | **P-value** | **Spearman CC** | **P-value** |
| --- | --- | --- | --- | --- |
| **Baseline MAGNIFI-CD Score** | 0.846 | <.001 | 0.835 | <.001 |
| **Follow-Up MAGNIFI-CD Score** | 0.861 | <.001 | 0.864 | <.001 |
| **Baseline MVAI Score** | 0.820 | <.001 | 0.823 | <.001 |
| **Follow-Up MVAI Score** | 0.854 | <.001 | 0.865 | <.001 |
| **Baseline PEMPAC Score** | 0.823 | <.001 | 0.809 | <.001 |
| **Follow-Up PEMPAC Score** | 0.837 | <.001 | 0.839 | <.001 |

Area under the curve analyses for MR scoring systems to assess a change in disease severity on the Likert severity score from baseline MR to follow-up MR, defined as a change of 2 or more units, are presented in **Table 6**. In patients who met the study definition criteria for change in disease severity (n=17), all scoring systems demonstrated an area under the curve of at least 0.93 (**Table 6**).

**Table 6.**
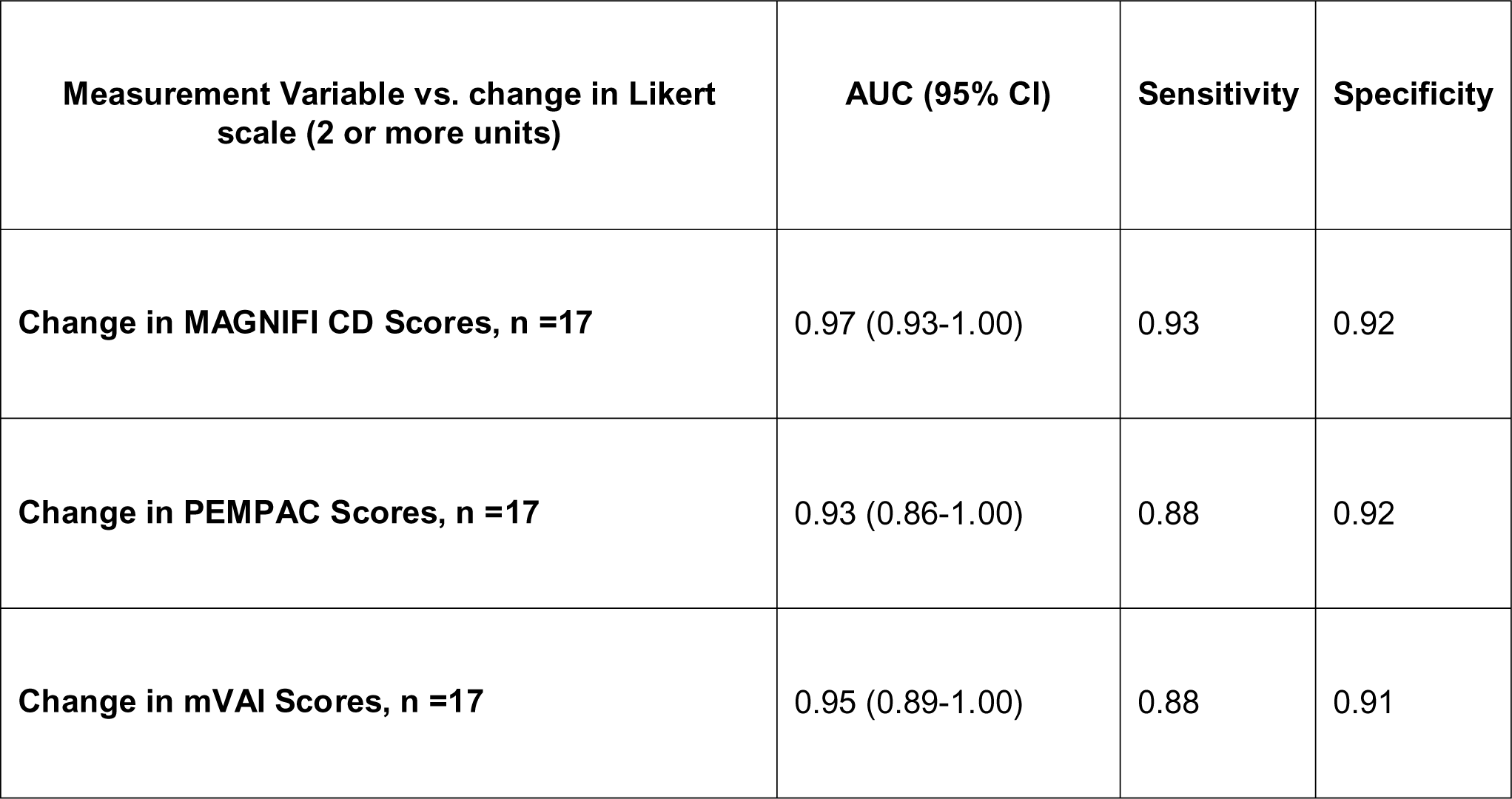
Responsiveness of MR imaging scoring systems to detect change in disease severity on the Likert scale.

| **Measurement Variable vs. change in Likert scale (2 or more units)** | **AUC (95% CI)** | **Sensitivity** | **Specificity** |
| --- | --- | --- | --- |
| **Change in MAGNIFI CD Scores, n =17** | 0.97 (0.93-1.00) | 0.93 | 0.92 |
| **Change in PEMPAC Scores, n =17** | 0.93 (0.86-1.00) | 0.88 | 0.92 |
| **Change in mVAI Scores, n =17** | 0.95 (0.89-1.00) | 0.88 | 0.91 |

Performance of the MR scoring systems compared to the study radiologists’ MR follow-up designation of markedly worsened, worsened, unchanged, improved, or markedly improved is presented in **Table 7**. **Figure 3** displays the top-ranked MR variable that influenced the study radiologists’ designation of Likert responsiveness at follow-up, as depicted in the heat map plot. The heat map illustrates the radiologists’ identification of the most influential MR factor for categorizing follow-up MR examinations as improved disease severity, stable, or worsened.

**Figure 3.**
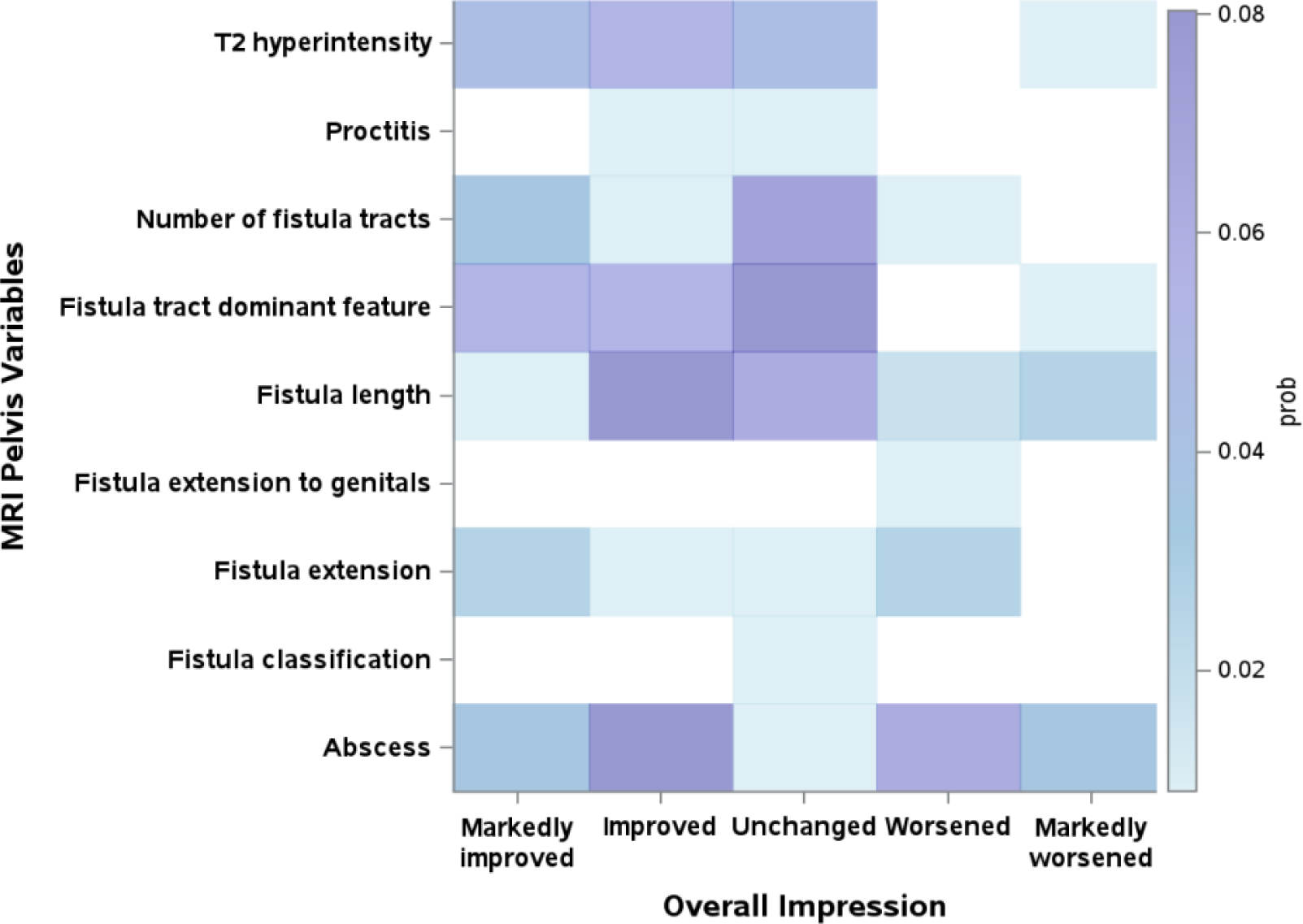
Heat map plot illustrating the study radiologists’ identification of the most influential MR factor for categorizing follow-up MR examinations as improved disease severity, stable, or worsened. Radiologists ranked their ‘top 5’ MR variable from 1 to 5 for each follow-up case, and the heat map displays the top-ranked variable for each follow-up MR. The colors represent the frequency of each variable ranked #1, with darker shades indicating higher rankings.

**Table 7.** Univariate association of MR scoring systems with study radiologists’ subjective overall impression of change from baseline MR to follow-up.

|  | Radiologists' subjective impressions |  |  |  |  | p-value* |
| --- | --- | --- | --- | --- | --- | --- |
|  | Markedly worsened | Worsened | Unchanged | Improved | Markedly improved |  |
| **MAGNIFI-CD, median (IQR), N = 85** | 8 (6, 12) | 2 (0, 3) | 0 (0, 0) | -2 (-7, -2) | -8 (-9, -4) | <.001 |
| **mVAI, median (IQR), n = 95** | 7 (3.5, 10.2) | 1.45 (0, 2.4) | 0 (0, 0) | -1.5 (-4.2, -0.5) | -3.9 (-7.5, -2.4) | <.001 |
| **PEMPAC, median (IQR), n = 96** | 15 (4, 17) | 0 (0, 2) | 0 (0, 0) | -1 (-13, 0) | -8 (-11, -3) | <.001 |

\*non-normal distribution of the data non-parametric Kruskal-Wallis test to calculate the p-value.

## Discussion

This study evaluated the radiologist performance characteristics of CD-PAF MR scoring systems, including MAGNIFI-CD [15], mVAI [14], and PEMPAC [16], in a real-world cohort of patients with CD-PAF who underwent serial imaging with pelvic MR. In our study design, the MR scoring systems are responsive in assessing changes in disease severity and show associations with radiologists’ subjective impressions. MR imaging using a fistula protocol is the current reference standard for assessing disease activity in CD-PAF [4–6]. However, the inconsistent agreement among the individual components of the CD-PAF MR scoring systems indicates a need for further refinement and standardization. The variability in certain components and the extensive number of reportable factors required for some scores challenge their integration into regular clinical interpretations.

CD-PAF patients have limited therapeutic options and reduced quality of life [5]. Despite medical treatment with biologics and surgical seton placement, remission is achieved in only a third of patients at 1 year, and many experience recurrence within 2 years due to poorly characterized treatment responses [5]. Given this clinical landscape, measuring improvement in perianal fistulas on MR has become increasingly integrated into CD-PAF clinical trials [21, 22]. An analysis of published and ongoing randomized trials in CD-PAF highlights the increasing incorporation of MR imaging into the primary endpoints. In 19 published randomized trials of CD-PAF ranging in publication year 1980 to 2020, 4 of 19 incorporated MR into the primary endpoint. These 4 trials represented 4 of 6 trials published from 2015 to 2020. Furthermore, the analysis of ongoing trials showed MR incorporated into the primary endpoint in 4 of 8 ongoing trials [21]. Similarly, a meta-analysis of published clinical trials for stem cell therapy in CD-PAF showed MR findings incorporated into the outcomes measures in 15 of 29 studies, with a similar trend of more recent published trials including MR more frequently [22].

This trend of incorporating MR imaging into CD-PAF clinical trials aligns with the increasing recognition of radiological healing as an important treatment goal [6–12]. However, there is no consensus on the definitive criteria for CD-PAF treatment response on MR imaging. A systematic review of CD-PAF clinical trials with imaging assessment reveals considerable variability in defining improvement or fistula closure by MR among published studies [23]. In a secondary analysis of a clinical trial involving 50 CD-PAF patients, van Rijn et al. [24] developed a novel degree of fibrosis scale for assessing the long-term closure of perianal fistulas on MR. This scale had the study radiologists subjectively estimate the fibrosis volume within the fistula tract using T2-weighted imaging, with scores ranging from 0% to 100%. They found that a completely fibrotic fistula (100% on the scale) robustly indicated long-term closure [24]. In the current study, the “dominant feature” of the primary tract, predominantly fibrous, was frequently observed on the heatmap (**Figure 3**) for both unchanged and improved follow-up cases as the most frequent study radiologists’ top choice for making their following designation.

Numerous scoring systems have been developed for MR of the pelvis, including the VAI [17], and the three scoring systems evaluated in this study, mVAI [14], MAGNIFI-CD [15], and PEMPAC [16]. PEMPAC was developed in a pediatric cohort of CD-PAF [16]; we incorporated this scoring system in this study to assess its performance in an adult cohort. These MR scoring systems have been incorporated into primary and secondary endpoints in CD-PAF clinical trials [21, 22]. Interobserver reliability is up to moderate for some of these scoring systems [23]. Each iteration of the scoring systems, from VAI, mVAI, to MAGNIFI-CD, has sought to improve assessment and definition of imaging improvement in CD-PAF. While interobserver reliability has improved with the most recent MR scoring system, MAGNIFI-CD [15], this is most frequently assessed at a CD-PAF patient’s MR baseline where the fistula is more severe than post-treatment.

Despite the limited agreement of the individual MR factors, the CD-PAF MR scoring systems exhibited strong and significant associations with radiologists’ subjective severity assessments and follow-up changes, aligning with their expected performance. Given that MAGNIFI-CD [15] and PEMPAC [16] were developed through a combination of expert opinion and assessment of disease severity on a 100 mm visual analog scale, the strong association between the CD-PAF MR scoring systems and radiologists’ subjective severity assessments is consistent with their design. However, the magnitude of change in these scores that would constitute radiological healing remains undefined without a direct correlation between pathology and imaging findings.

One limitation of the study is that clinical outcomes were not reported in this study or compared to the scoring systems. Additionally, the scans were performed over a study period of 10 years, and changes in MR scanners and protocols may have impacted the study findings. Additionally, the scans were performed at varying intervals that resemble real-world practice more than set intervals, as seen in CD-PAF clinical trials. MAGNIFI-CD [15] was developed in an imaging dataset that incorporated specific fistula anatomy due to inclusion/exclusion criteria of the ADMIRE-CD study [25–27], different from the broader fistula anatomy imaged in this study. Similarly, the PEMPAC index [16] was developed in a pediatric cohort, and this study constitutes its application in an adult cohort. While some MR scoring systems have shown moderate inter-reader agreement, our study, similar to other systematic reviews of CD-PAF clinical trials with imaging assessment [23], found considerable variability in predicting responders and non-responders, with some studies not achieving a significant improvement in post-treatment MR scores. These inconsistencies highlight the need for further refinement and standardization of MR scoring systems in assessing treatment response for CD-PAF.

In conclusion, MR scoring systems for CD-PAF, including MAGNIFI-CD, mVAI, and PEMPAC, demonstrate moderate inter-reader agreement but show variability when correlating with radiologic improvement. While they strongly align with radiologists’ subjective assessments of severity and treatment response, the inconsistencies among their individual components suggest a need for further refinement. The current variability restricts the adoption of these systems in clinical interpretations and their recommendation as endpoints in CD-PAF clinical trials. Future research should prioritize reaching a consensus on CD-PAF treatment response criteria using MR imaging and refining these systems to guide treatment decisions and improve patient outcomes.

## Data Availability

All data produced in the present study are available upon reasonable request to the authors.

